# Is Multimodal Better? A Systematic Review of Multimodal *versus* Unimodal Machine Learning in Clinical Decision-Making

**DOI:** 10.1101/2025.03.12.25322656

**Authors:** Alaedine Benani, Stéphane Ohayon, Fewa Laleye, Pierre Bauvin, Emmanuel Messas, Sylvain Bodard, Xavier Tannier

## Abstract

Machine learning has demonstrated success in clinical decision-making, yet the added value of multimodal approaches over unimodal models remains unclear. This systematic review evaluates studies comparing multimodal and unimodal ML algorithms for diagnosis, prognosis, or prescription. A comprehensive search of MEDLINE up to January 2025 identified 97 studies across 12 medical specialties, with oncology being the most represented. The most common data fusion involved tabular data and images (67%). A risk of bias assessment using PROBAST revealed that 57% of studies had a low risk of bias, while 41% had a high risk. Multimodality outperformed unimodality in 91% cases. No correlation between dataset sample size and added performance has been observed. However, considerable methodological heterogeneity and potential publication bias warrant caution in interpretation. Further research is needed to refine evaluation metrics and hybrid model architectures based on specific clinical tasks.

**MeSH Terms:** Humans [B01.050.150.900.649.313.988.400.112.400.400], Machine Learning [L01.224.050.375.530], Clinical Decision-Making [E01.055], Systematic Review [V03.850].

## Introduction

Artificial intelligence (AI) has emerged as a transformative technology in healthcare, offering opportunities to improve clinical decision-making and therefore patient outcomes. By leveraging machine learning (ML)—a subset of AI—healthcare systems can analyze large-scale and heterogeneous datasets to develop algorithms that support diagnosis, prognosis, and treatment planning^1^. AI-driven models have already demonstrated significant success in fields such as radiology^2^, dermatology^3^ and pathology^4^, where they assist clinicians in interpreting medical images, predicting disease progression, and planning treatment strategies^5^.

Traditional machine learning models often rely on unimodal data, which refers to a single type of input, such as medical imaging (e.g., Computed Tomography [CT], Magnetic Resonance Imaging [MRI], ultrasound [US]), structured data (e.g., laboratory results), unstructured data (e.g., clinical notes, free text), signal (e.g., electrocardiogram [EKG]), videos data or omics (e.g., genomics, transcriptomics). While unimodal models have yielded notable achievements, they are suspected to be inherently limited in their ability to provide a comprehensive understanding of a patient’s condition^6^. For example, predicting future ischemic cardiac events with only clinical tabular data may reveal some insights (as age and LDL-cholesterol are widely recognized risk factor) but it lacks critical information from genetic (cardiovascular PRS), physiological (EKG), imaging (coronary arteries evaluations) to provide an informed prediction - adding EKG and coronary imaging might enhance ischemic cardiac events prediction. This fragmented approach might restrict the predictive performance and generalization of ML models in diverse healthcare settings^7^. Multimodal data, on the other hand, integrates information from multiple data types. This integrative approach allows AI systems to capture complementary features across different modalities, enabling a more exhaustive representation of the clinical task to address.

Despite its potential, the use of multimodal data in healthcare applications remains underdeveloped due to several challenges^5^. Current unimodal models dominate the landscape, primarily because integrating heterogeneous data types poses methodological and technical barriers. These include difficulty in data accessibility, differences in data formats, challenges in feature alignment, inconsistent data quality across the various datasets, preprocessing requirements that differ between categories, biases that are harder to identify and reduce, more complex validation methods^8^. Additionally, multimodal approaches require higher computational resources (in terms of computing power, active memory capacity, and storage capacity) and more sophisticated algorithms to process and analyze high-dimensional dataset, capable of capturing complex interactions while avoiding overfitting or divergences.

The reliance on unimodal data underscores a potential gap in healthcare AI, as it might limit the ability to fully capture the complexity of clinical information. In recent years, numerous studies have investigated multimodal algorithms, often comparing their performance to unimodal approaches. Despite this growing body of research, the practical relevance of these approaches, the rigor of the methodologies employed, and the suitability of the proposed use cases remain inadequately understood.

However, while multimodal machine learning has gained increasing attention, there remains a lack of comprehensive synthesis regarding its actual benefits and limitations in clinical practice. To bridge this gap, this systematic review aims primarily to address the following research question: “In which cases the use of multimodal data enhances the performance of machine learning algorithms compared to unimodal data in clinical decision-making tasks?”

The objective of this question is to determine whether the integration of multiple data modalities enhances the performance and generalizability of machine learning models across a range of clinical decision-making tasks and, if so, in which settings.

Therefore, there are three secondary objectives to this study.

**The first** one is to identify the types of multimodal data commonly utilized in multimodal settings. This includes characterizing the combinations of data modalities— tabular clinical data, unstructured free text, medical imaging, signal data, videos data, and omics data—employed in machine learning models. Particular attention will be paid to understanding which combinations are most prevalent, in which clinical contexts they are applied, and sample size of the datasets used.

**The second** one is to assess the methodological approaches used in integrating multimodal data for algorithm development. This involves evaluating how multimodal data are preprocessed, combined, and incorporated into machine learning models. Specific methodological aspects, including the use of classic machine learning or advanced deep learning methods, the integration of multimodal data through fusion methods (combining information from different modalities at input, intermediate, or output stages), and evaluation frameworks, will be analyzed to identify trends and best practices in the field.

**The third** and last one is to examine the benefits, limitations, and challenges associated with using multimodal data in healthcare settings. This objective focuses on elucidating the added value of multimodal data integration in terms of improving predictive performance and clinical utility. Additionally, the review will highlight potential bias, quantity of the open sourced, FDA or CE cleared algorithms and the mention of a medico-economic evaluation.

## Results

### Study selection

We identified 352 records through database search (**Supplementary Table 1**). After removing 2 duplicates (because MEDLINE includes medXriv and some arxiv journals even when they are published in peer-reviewed journals subsequently), 350 unique records remained and were screened for titles and abstracts. Of these, 134 full-text articles were assessed for full-text eligibility. 37 articles were excluded, with reasons detailed in **Figure 1**, mainly absence of multimodal versus unimodal comparison (n = 28). One article might have been excluded for multiple reasons. Non-peer reviewed articles were excluded. Ultimately, 97 studies met all the inclusion criteria and were included in the review. The study selection process is summarized in the PRISMA flow chart (**Figure 1**).

**Figure 1:**
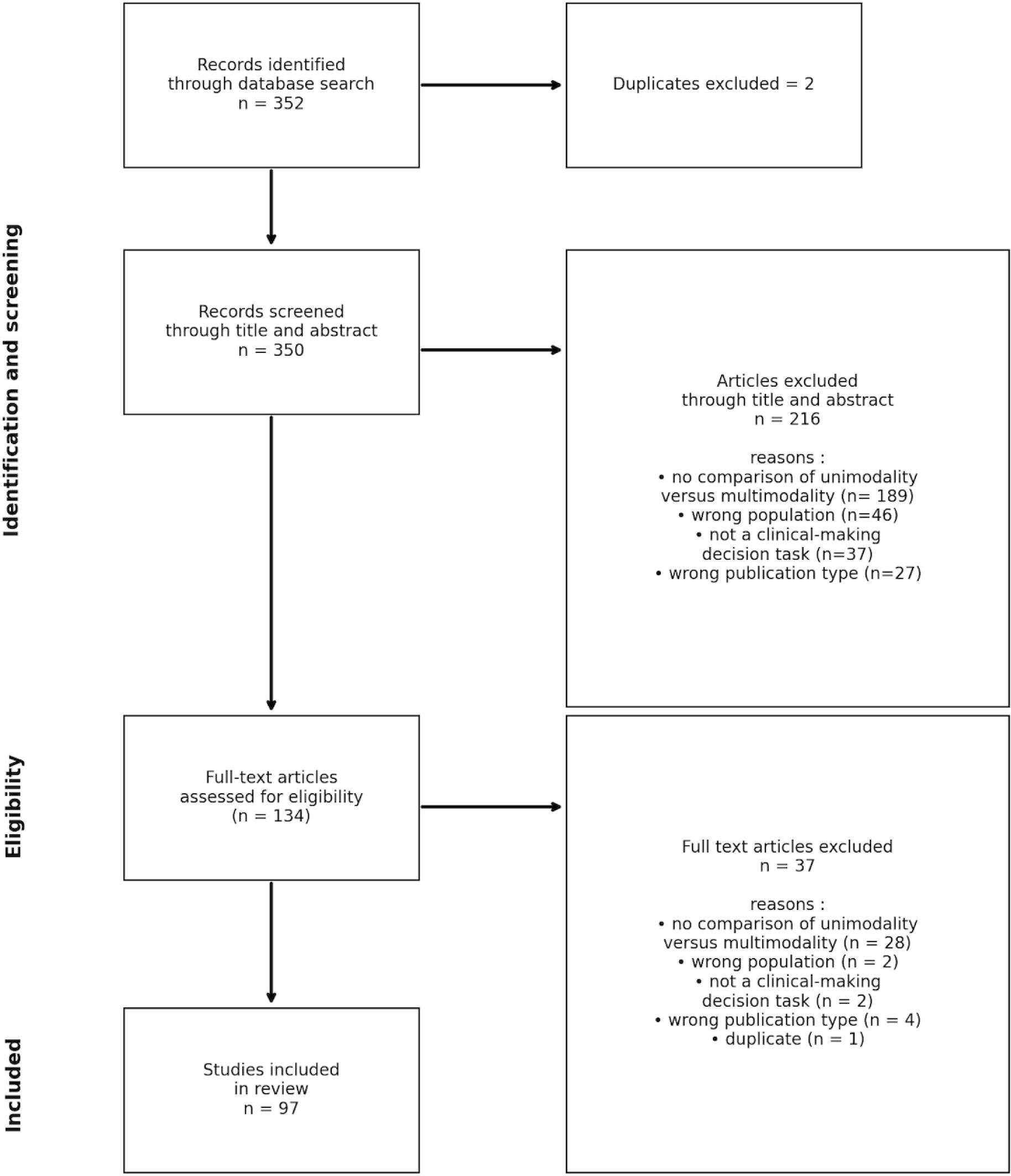
PRISMA flowchart. Visual representation of the search strategy, data screening and selection process of this systematic review.

### Characteristics of Included Studies

Regarding the research setting, 51 studies (53%) were multicenter studies, 39 studies (40%) were conducted within a single institution, and 9 studies (9%) were bicentric studies (**Supplementary Table 2**).

In terms of publication year, 9 studies were published in 2020 or earlier, with a continuous increase in the number of publications in subsequent years : 10 studies in 2021, 12 in 2022, 22 in 2023, 36 articles in 2024. In January 2025 only, there have been as many articles (10, 10%) as the whole 2021 year (**Supplementary Table 3**).

Geographic distribution showed that 30 studies (31%) originated from the United States, 26 studies (27%) from China, and 20 studies (21%) from Europe. Additionally, 11 studies (11%) used datasets from mixed international sources (**Supplementary Figure 1**).

The medical specialties addressed were diverse (**Supplementary Figure 2**). Cancer was the most represented specialty, comprising 35 studies, followed by cardiovascular diseases with 14 studies, and neurology with 13 studies.

The types of multimodal data utilized were greatly predominantly combinations of tabular data and imaging, reported in 65 studies. Combinations of omics data and imaging were used in 8 studies. **Figure 2** describes the most frequent combination of data types.

**Figure 2:**
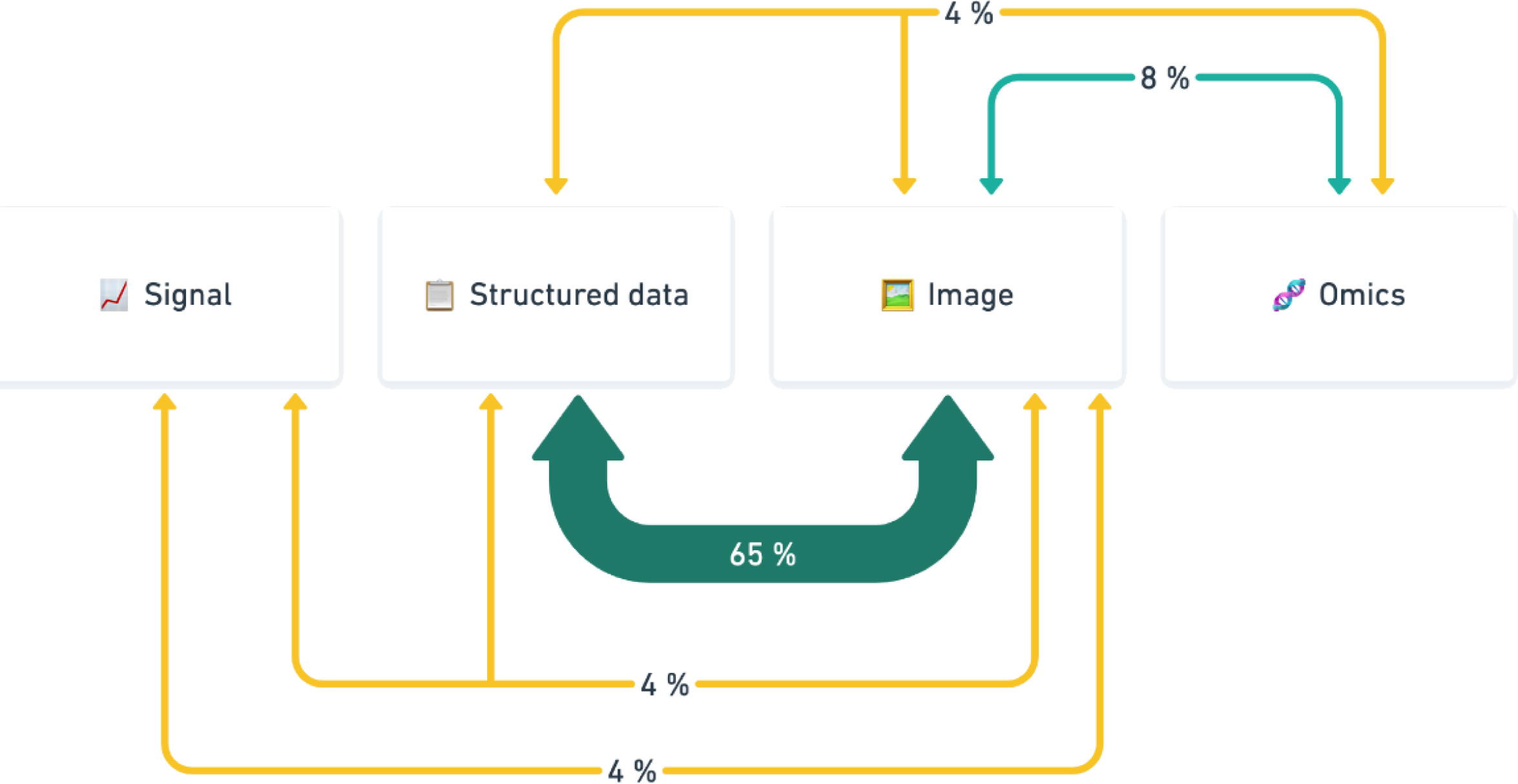
Combination of modalities across all studies. Distribution of modality combinations across the studies included in this systematic review. The four primary data modalities—Signal, Structured Data, Image, and Omics—are depicted, along with the frequency of their co-occurrence in multimodal machine learning models for clinical decision-making tasks. Only modalities with ≥ 4% of combinations are displayed.

The tasks of the algorithms included on this systematic review were categorized as follows : diagnosis (identification of a current disease), prognosis (prediction of future disease apparition, or worsening, or future events), prescription.

Prognosis tasks were the focus of 50 studies, while 49 addressed diagnosis tasks (**Supplementary Figure 3**). Interestingly, no studies included targeted prescription tasks.

Of note, only one article used a generative AI model (ChatGPT-4V)^9^. Key characteristics of the included studies are summarized in **Table 1**.

**Table 1:**
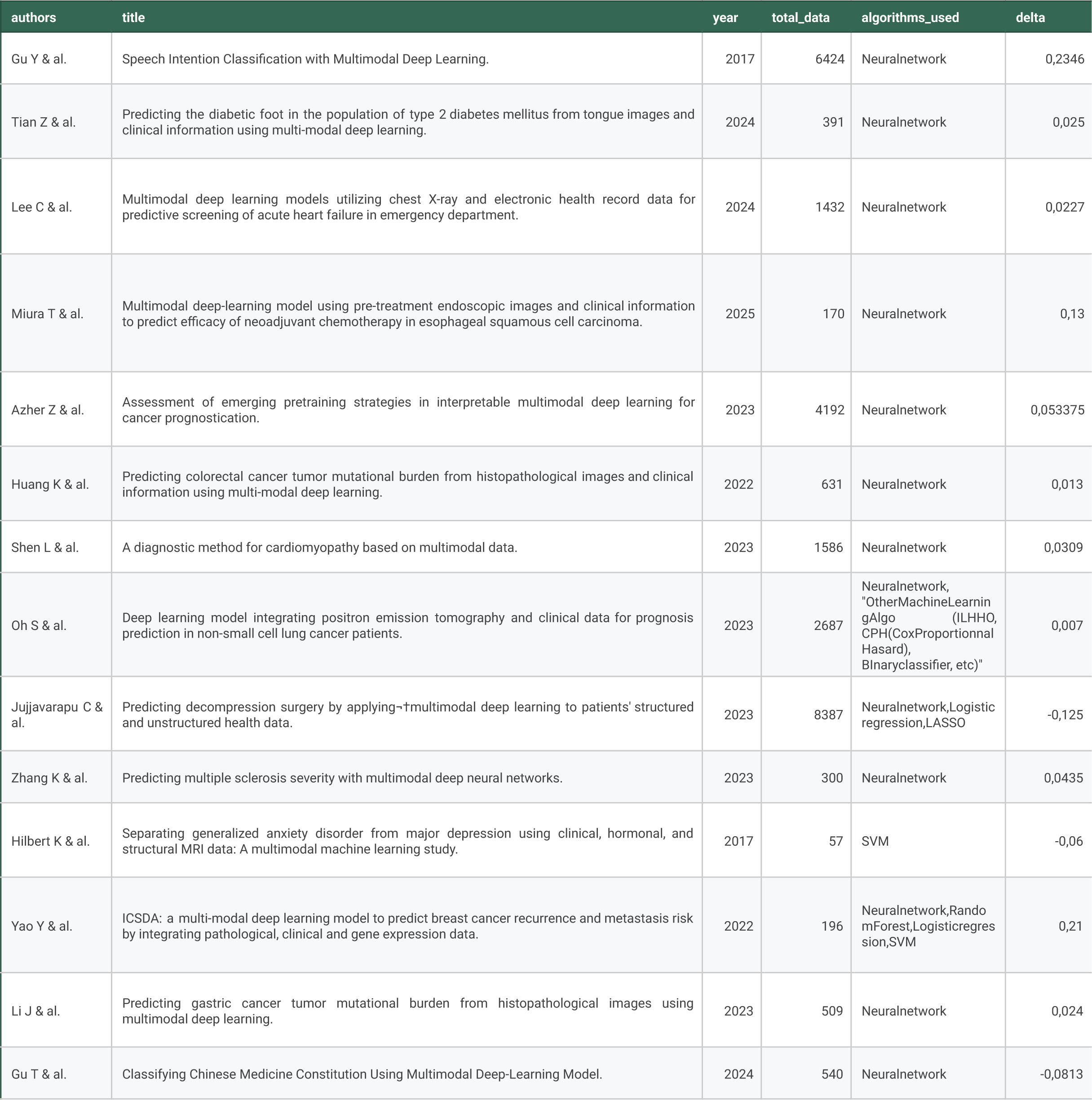
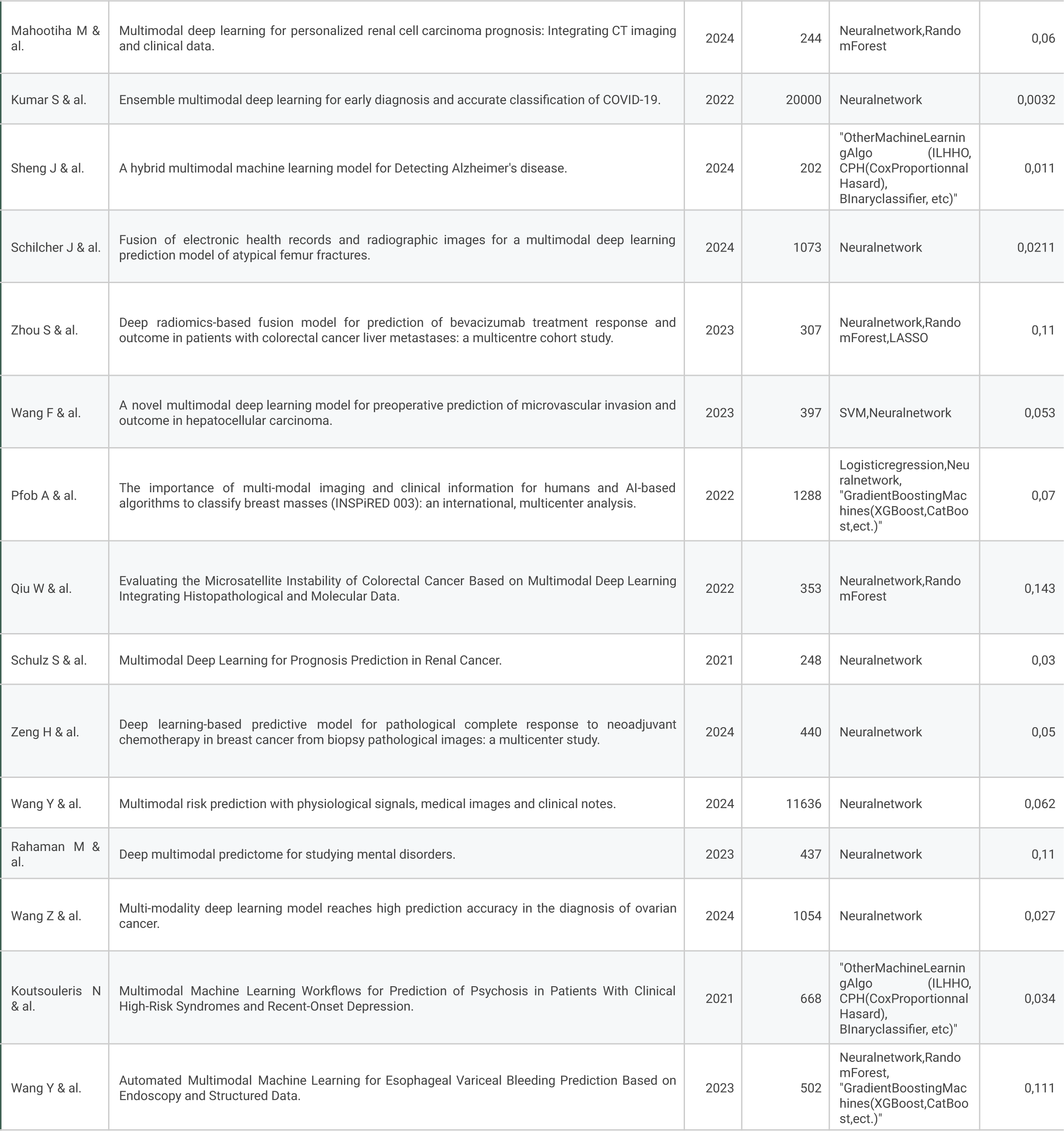
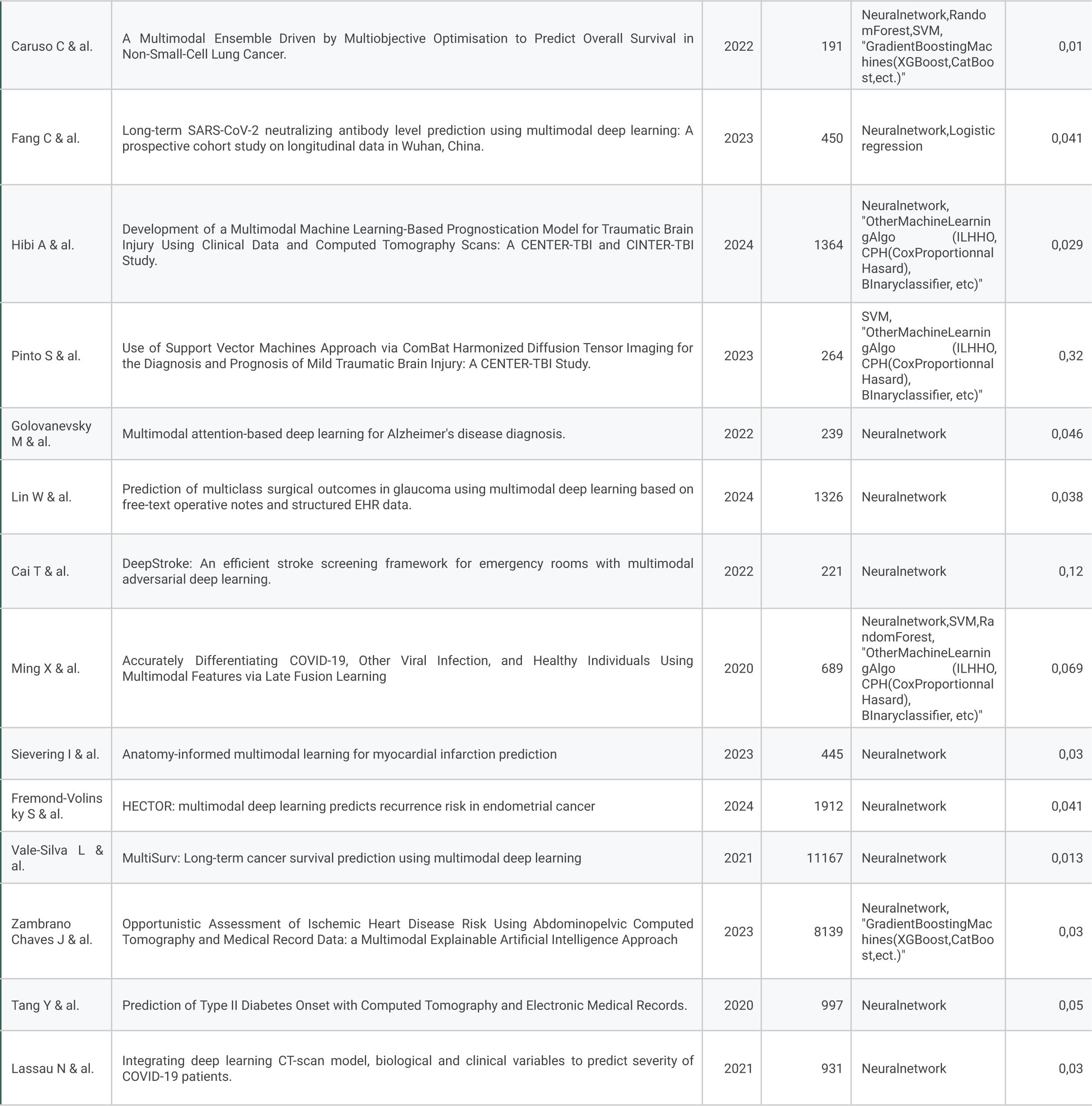
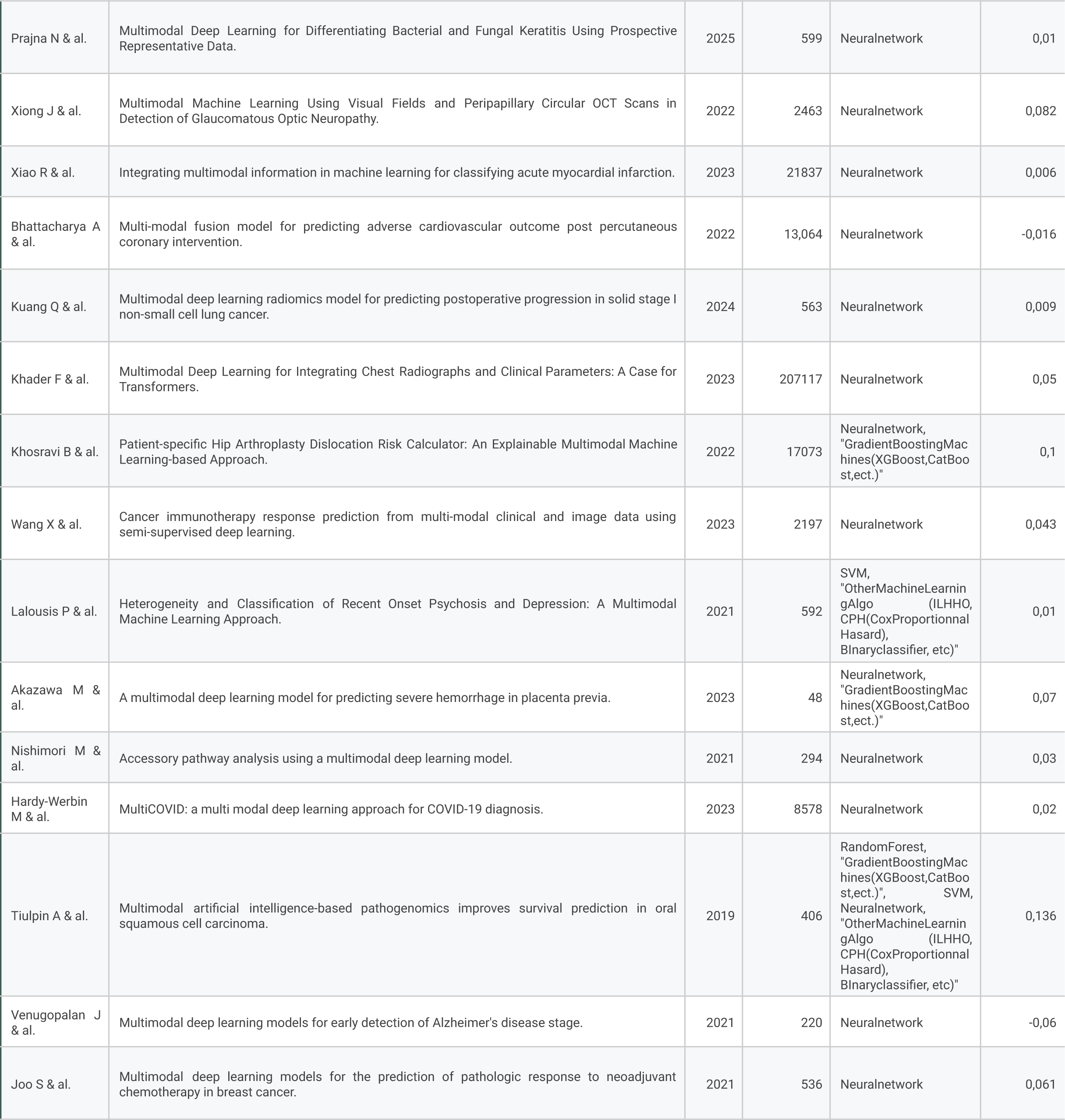
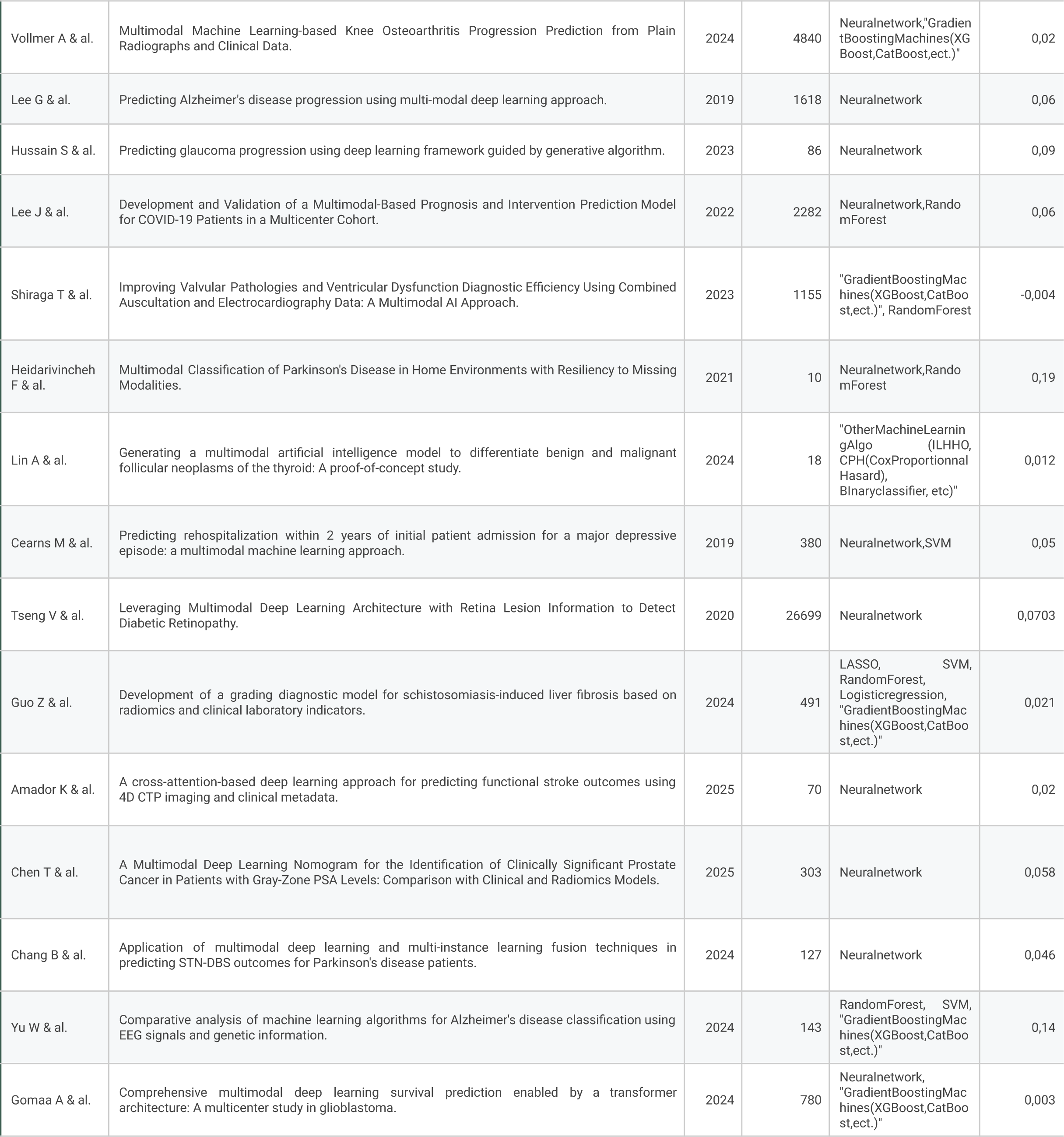
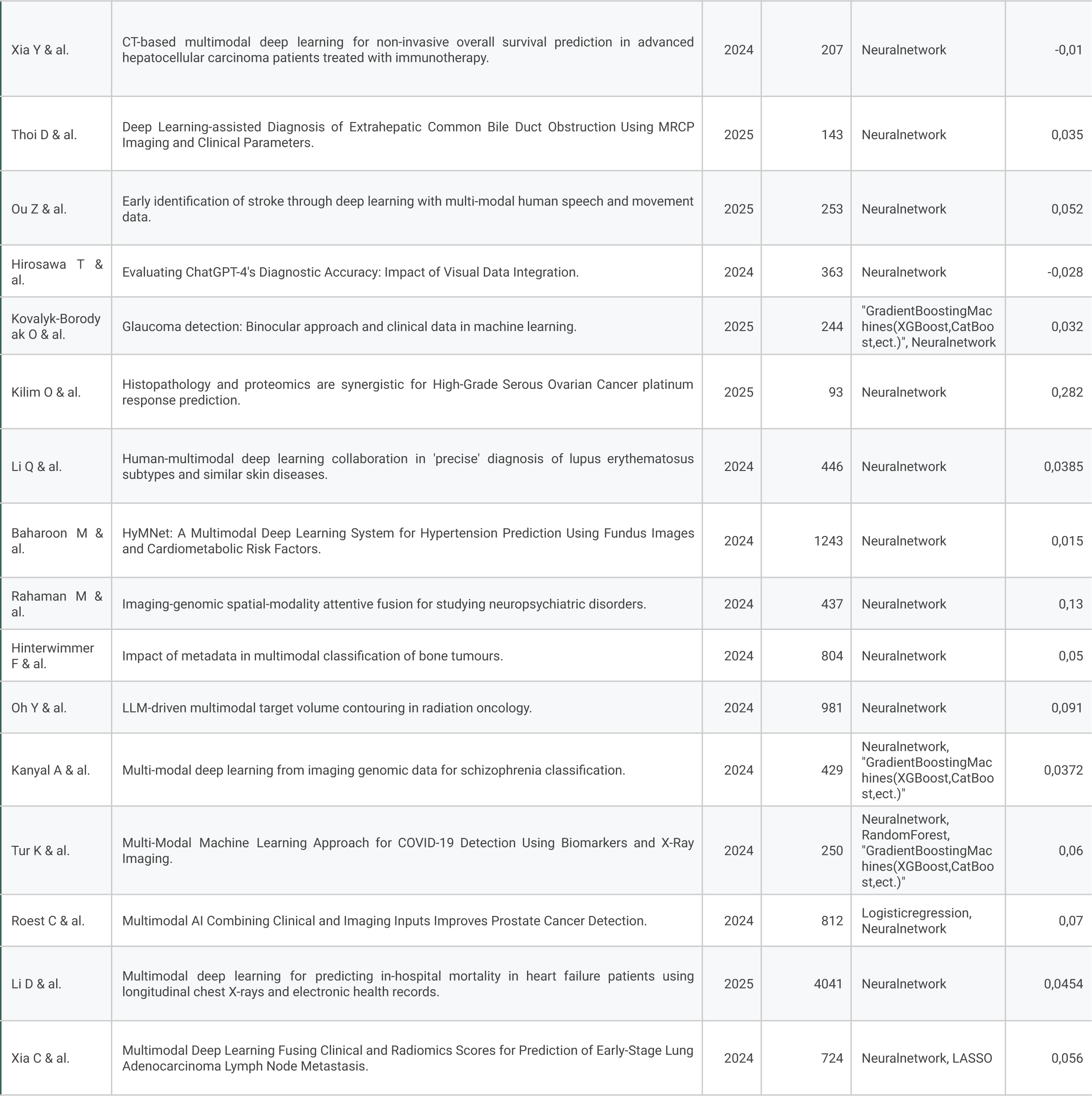
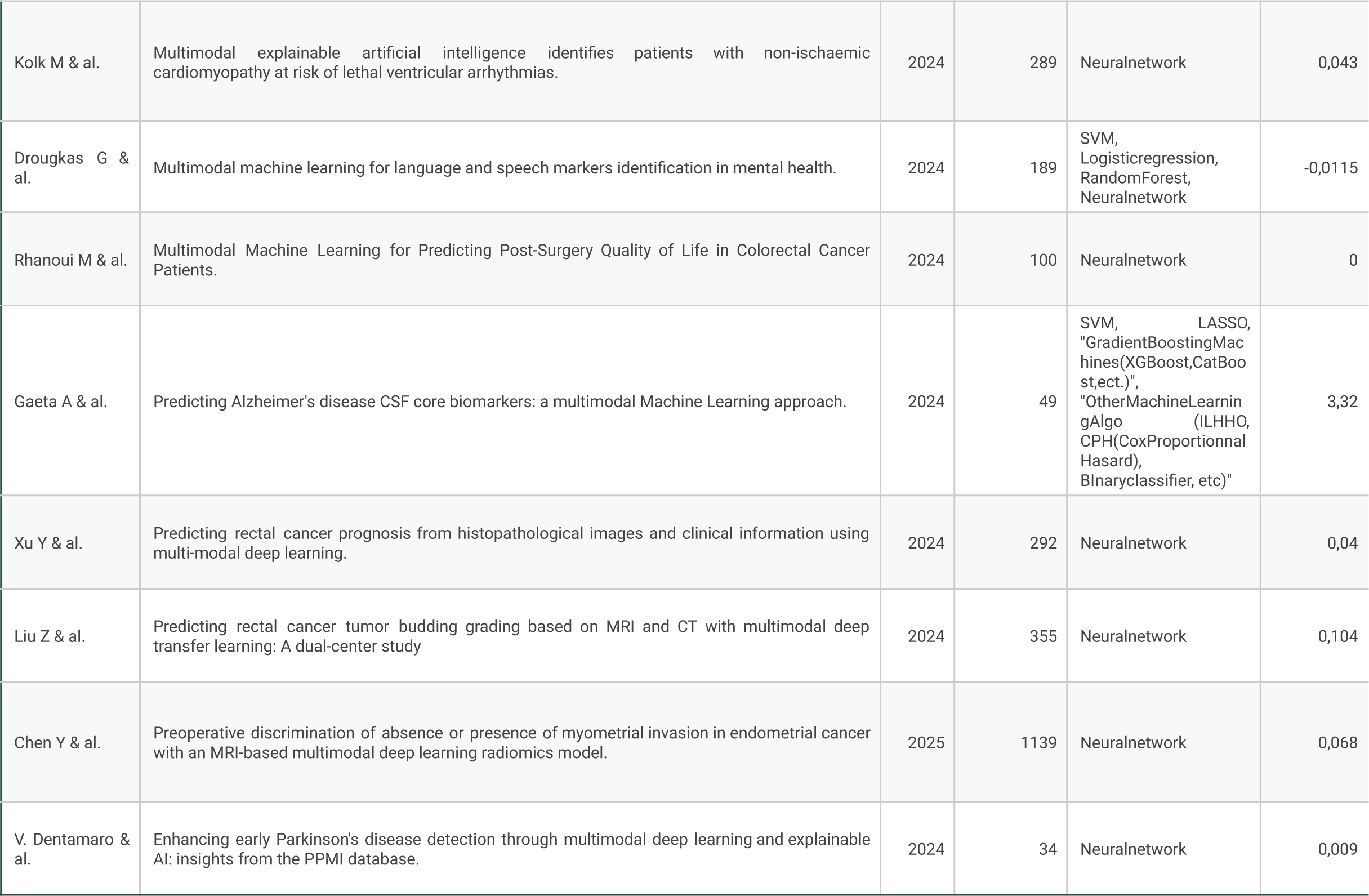
Key characteristics of included studies.

### Performance Comparison

The included studies exhibited considerable variation in methodologies, including the types of classification algorithms employed, the methods used for algorithm training, the features utilized, and the evaluation metrics reported. Despite this heterogeneity, common trends emerged in the performance evaluation of multimodal versus unimodal approaches.

The most frequently emphasized evaluation metrics across studies were the area under the receiver operating characteristic curve (AUC), reported in 66 (68%) of studies, followed by accuracy (20 studies, 21%) and the concordance index (C-index) in 8 (8%) studies (**Supplementary Figure 4**). Of note, most of the studies reported many evaluation metrics, but one was almost always emphasized.

Of the 97 studies included in the review, 88 studies (91%) demonstrated that multimodal approaches outperformed unimodal algorithms (**Figure 3**).

**Figure 3:**
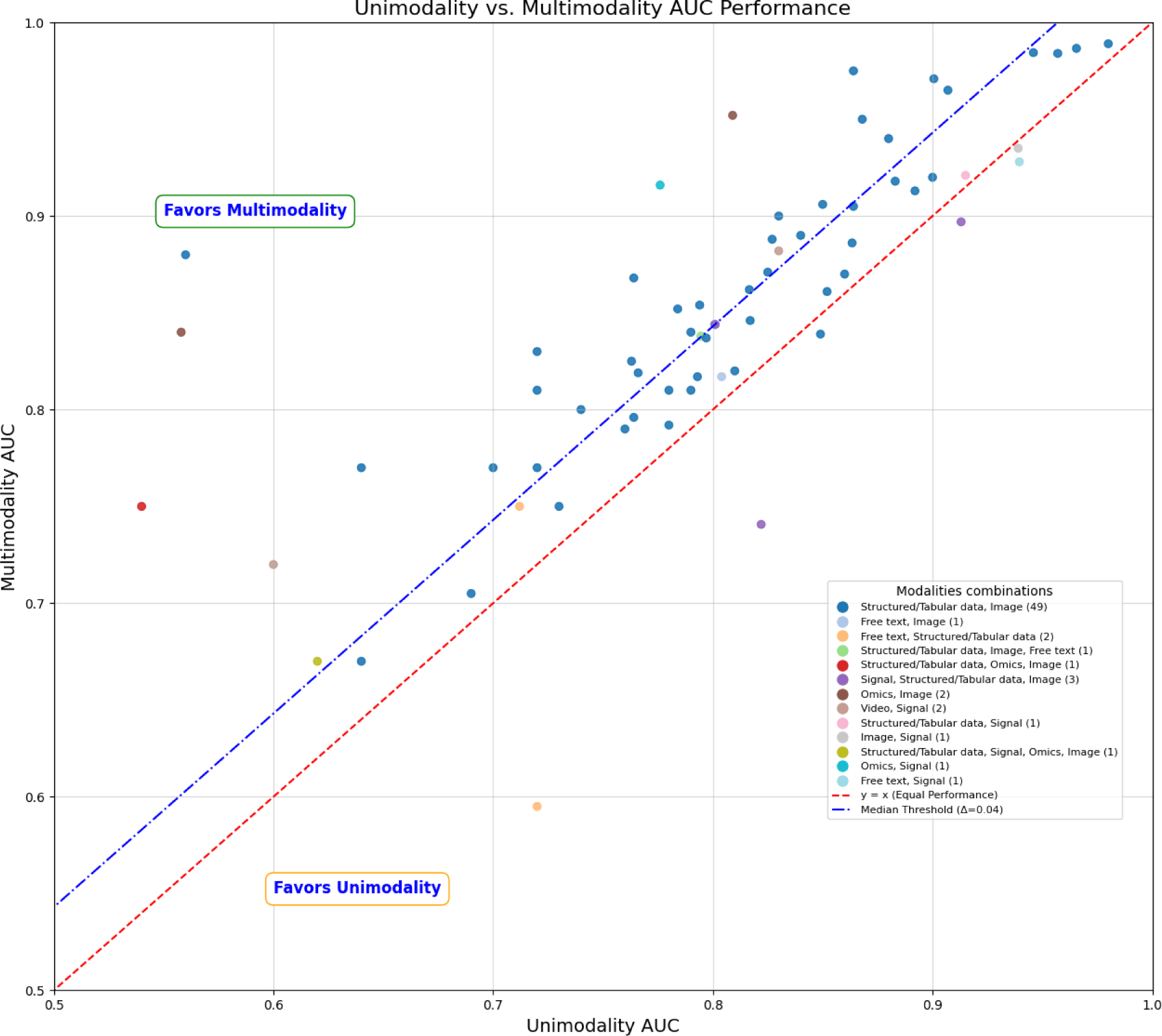
AUC added performance through multimodal approach across all modalities. This scatter plot illustrates the comparative performance of unimodal and multimodal machine learning models across studies included in this systematic review. Each point represents an individual study, with the unimodal area under the curve (AUC) score on the x-axis and the corresponding multimodal AUC on the y-axis. Points above the diagonal red dashed line (y = x) indicate studies where multimodality outperformed unimodality, whereas points below this line correspond to cases where unimodal models demonstrated superior or equivalent performance. The blue dashed line represents the median performance threshold (Δ = 0.04), capturing the general improvement observed when integrating multiple data modalities. The color of each data point corresponds to a specific modality combination, as detailed in the legend. Structured/tabular data combined with imaging is the most frequent multimodal approach, followed by other combinations, including free text, signal, and omics data.

To quantify the added performance due to multimodality, we calculated the performance delta, defined as the difference between the best-performing multimodal and unimodal algorithms in each study. Across the included studies, the performance deltas ranged from a minimum of -0.13 to a maximum of 0.32. The median of added performance is 0.04. This distribution of AUC performance improvement for 66 studies is illustrated in **Figure 3**, emphasizing that while most studies reported improved AUC, 26 (50%) of them reported a gain inferior to 0.04. A subset (10 studies) achieved substantial enhancements (above 0.10), with a few (6 studies) reporting negative improvements.

The combination of free text and signal data yielded the greatest improvements, highlighting the value of integrating these complementary modalities to enhance algorithmic performance.

To further explore the underlying reasons for the performance gains observed with multimodal approaches, we tested the hypothesis that the magnitude of added performance is directly related to the medical relevance of the added modality. Specifically, the added modality in each study was classified into three groups based on its suspected contribution to the clinical decision-making process. The first group (High) included modalities deemed to provide entirely new and medically meaningful information, unavailable in the initial modality. The second group (Moderate) comprised modalities that were suspected to provide overlapping information with the initial modality, while the third group (Low) consisted of modalities that did not appear to contribute additional relevant information. If many modalities were added, the most relevant group was kept for each article. If the added value was uncertain, the ‘Uncertain’ label was attached to it.

Across the 66 studies with AUC performance, the distribution of added modalities was as follows: 43 (65%) were classified as providing entirely new information (high meaning, e.g., fusing genetics information with imaging), 21 (32%) as overlapping with existing information (moderate meaning, e.g., fusing clinical parameters with imaging), and 2 (3%) as offering minimal additional information (low meaning, e.g., fusing echocardiogram with ECG while ECG signal appears on the echocardiogram slides). Interestingly, all the articles that had a low meaning added modality were found to have a limited effect on the multimodal performance, below the median (0.04) of added performance (**Supplementary Table 4**). However, probably due to the very low sample of the low-meaning additional information, we did not observe a significant correlation between the medical meaningfulness of the added modality and the improvement in performance of the multimodal approach compared to the unimodal approach. Of note, most of the studies with a Moderate added values were combining tabular and images data, as the informations in one modality are frequently overlapping to some extent with the information on the other modality. Therefore, in this study, clinical significance appears as a necessary but insufficient criteria to have a significant added value of a modality.

## Methodological Approaches

### Dataset sizes

The 97 included studies used datasets of widely varying sizes, reflecting the diversity of modalities and data sources. The smallest dataset consisted of 10 samples, while the largest dataset contained 207,117 samples, and the median at 631 samples. The distribution of dataset sizes is illustrated in **Figure 4**. We did not observe any <u>strong correlation</u> between the data size and the overall performance of the algorithm (Pearson coefficient *r* = 0.10). Also, we did not find <u>any correlation</u> between the dataset sizes and the added performance of the multimodal approach (Pearson coefficient *r* = -0.02).

**Figure 4:**
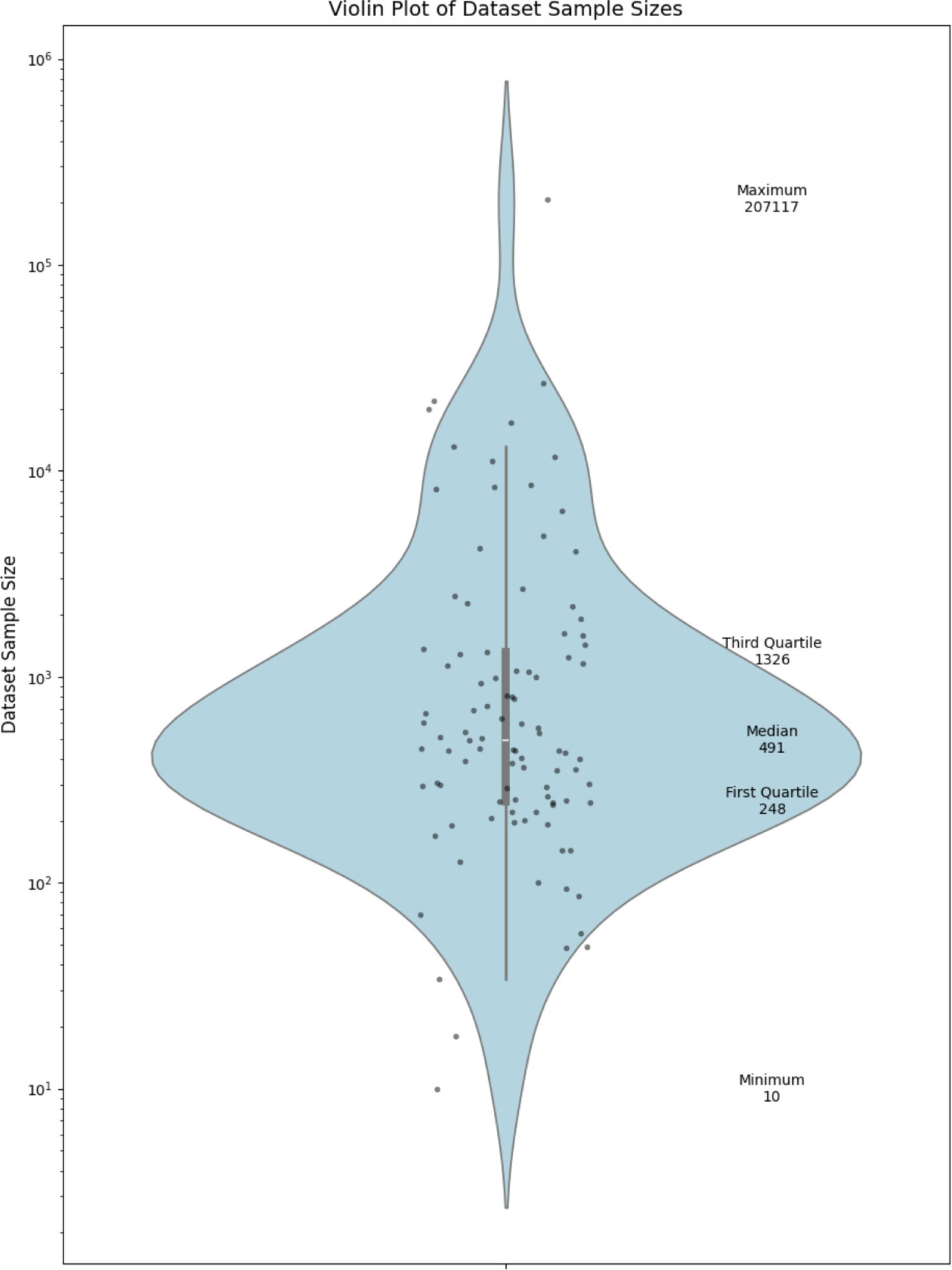
Violin-Plot visualization of datasets sample size. This violin plot illustrates the distribution of dataset sample sizes among the studies included in this systematic review. The y-axis is displayed on a logarithmic scale to accommodate the wide range of sample sizes. The box represents the interquartile range (IQR), with the lower and upper edges corresponding to the first quartile (244) and third quartile (1,288), respectively. The red line within the box denotes the median sample size (450).

### Evaluation methods

The evaluation methodologies employed across the 97 included studies were categorized into three distinct validation strategies, ranked in order of validation rigor^10^: split-sample validation, repeated split-sample validation, and validation using an external dataset. Split-sample validation, the least robust method, refers to a single random train-test split without the inclusion of a separate validation dataset. This strategy was utilized in 19 studies. Repeated split-sample validation, which encompasses cross-validation or bootstrap approaches but similarly lacks a dedicated external validation dataset, was the most commonly employed methodology, used in 45 studies. The most rigorous approach, validation through the use of an external dataset, was implemented in 32 studies. This method provides stronger evidence of generalizability by testing model performance on independent data not used during training or internal validation.

### Used algorithms

Across the 97 included studies, we classified the algorithms utilized into 7 types : Neural Networks (NN), Logistic Regression, LASSO, Boosting (XGBoost, GBM, CatBoost, and Adaboost), Random Forest, SVM and others (e.g., K-Nearest Neighbords, Cox Proportional-Hazards models). We chose to combine all the NNs into one category. This choice to group the NN category is motivated by the fact that there was a high diversity in the kind of NN used. The fully granular list of algorithms used in the papers is available in **Supplementary table 1**. In the rest of the paper, non-NN algorithms will be referred to as “ML Algorithms”. A total of 153 algorithms were used, classified as previously described. NN were the most frequently employed, representing 87 algorithms (57%) across the studies. Regression-based methods, including logistic regression and LASSO, accounted respectively for 7 and 5 algorithms (5% and 3%). Random forest models and boosting algorithms, such as XGBoost, gradient boosting machines (GBM), CatBoost, and AdaBoost, were each used in 15 algorithms and 16 algorithms (10%). Support vector machines (SVM) were implemented in 13 algorithms (9%), and 10 algorithms (7%) fell into other methodological categories. The distribution of algorithms employed in the included studies is summarized in **Table 2**.

**Table 2:** Categories of used algorithms.

| Algorithm Categories | Number of studies | Percentage |
| --- | --- | --- |
| Neural Network (NN) | 87 | 56,86% |
| Random Forest | 15 | 9,80% |
| SVM | 13 | 8,50% |
| LASSO | 5 | 3,27% |
| Logistic Regression | 7 | 4,58% |
| Gradient Boosting | 16 | 10,46% |
| Other | 10 | 6,54% |
| Total | 153 | 100,00% |

A comparative analysis of the algorithms employed in multimodal and unimodal settings showed that, in the majority of cases (79%), the same algorithms were utilized across both settings, with adjustments made only to accommodate the respective data modalities. Within the neural network (NN) category, variations were frequently observed in the specific type of network implemented. For instance, in a given article, the unimodal setting would use ResNet architectures, while VGG architectures would be used in the multimodal settings.

### Fusion methods

The included studies employed a range of fusion methods to integrate multimodal data, with varying prevalence across the different approaches. Early fusion, in which features from different modalities are combined at the input stage before model training, was reported in 5 studies. Hybrid fusion, also referred to as midfusion, which combines features at intermediate layers within the model, was utilized in 20 studies. Late fusion, the most common approach, was employed in 64 studies and involves combining outputs from separate unimodal models at the decision-making stage. In 8 studies, the fusion method was not explicitly stated in the main text, making it unclear how the integration of multimodal data was achieved. It is noteworthy that several studies explored multiple fusion methods within their analyses, often presenting results for numerous of the approaches tested. For consistency and clarity in this systematic review, we selected the fusion method reported as achieving the best performance or explicitly highlighted as the primary approach in each article.

### Other characteristics

The open-source status of the algorithms described in the included studies was assessed based on the availability of code through public repositories and the presence of associated open-source licenses. Of the 97 articles reviewed, 10 articles provided open-source code, primarily hosted on repository platforms such as GitHub. The licenses used for these repositories were diverse. A majority of the articles (n = 87) did not provide open-source code.

The review assessed whether the algorithms described in the included studies had received clearance as medical devices by regulatory bodies such as the U.S. Food and Drug Administration (FDA) or European Conformity (CE). Only one article (1.59%) referenced the use of an algorithm that was partially cleared as a medical device by both the FDA and CE. The remaining articles did not mention any medical device clearance.

We also assessed whether the included studies provided or referenced a medico-economic evaluation of the algorithms being developed or tested. Medico-economic evaluations are critical for understanding the cost-effectiveness and potential economic impact of integrating such algorithms into clinical workflows. None of the included articles provided or mentioned a medico-economic evaluation.

### Appraisal of methodological quality

A risk of bias assessment was conducted for all included articles using the PROBAST^11^ (Prediction model Risk of Bias ASsessment Tool) framework. This evaluation revealed that 41% exhibited a high risk of bias, 56% had a low risk of bias, and 2% were categorized as having an uncertain risk of bias (**Table 3**).

**Table 3:** Proportion of Risk of Bias across studies.

| Risk of bias | Number of studies | Percentage |
| --- | --- | --- |
| Low | 55 | 56,70% |
| High | 40 | 41,24% |
| Uncertain | 2 | 2,06% |
| Total | 97 | 100,00% |

The PROBAST framework evaluates bias across four domains: participants, predictors, outcomes, and analysis. Among these, the most frequent source of bias was identified in the “analysis” domain, with 39 studies classified as high risk of bias in this category. Notably, the overfitting risk emerged as the most common issue. This is explained by the amount of studies that do not use an external dataset for validation. Interestingly, 49 studies do not explicit their method for handling missing data.

Using AMSTAR-2^12^ (A MeaSurement Tool to Assess Systematic Reviews), we rated the overall confidence in the results as moderate, mainly due to our subject that is not concerned by randomization studies.

## Discussion

This systematic review evaluated 97 studies comparing unimodal and multimodal algorithms for clinical decision-making tasks across diverse medical specialties, dataset sizes, data modalities, and fusion methods. In 91% of the studies, multimodal approaches demonstrated superior performance compared to their unimodal counterparts. This consistent advantage highlights the strong potential of multimodal artificial intelligence in the training of learning algorithms in many healthcare use cases.

The recent emergence and application of multimodal methods reflect the advancements in the field^13^. Almost all the studies reviewed were published within the past five years, coinciding with the rapid evolution of computational frameworks, improved hardware capabilities, and declining costs of computation, memory, and data storage^14^. These developments, combined with the growing availability of biomedical data from large biobanks, electronic health records, medical imaging, wearable devices, and ambient biosensors, have created a fertile environment for the training and testing of multimodal AI algorithms^15^.

Our work highlights the predominance of neural networks, including advanced architectures such as GAN^16^, transformers^17^ and cross-attention mechanisms, further emphasizes the field’s technological maturation. These architectures are particularly well-suited for processing heterogeneous data sources, enabling the extraction of complementary features from multiple modalities.

As expected, structured data emerged as the most prevalent data type across the reviewed studies, with the combination of structured data and medical imaging being the most common (**Figure 2**). This prevalence likely reflects the relative ease of collecting and standardizing these data types in clinical settings^18^. However, with advancements in large language models (LLMs), the utilization of free text is expected to increase significantly in the coming years. Clinical free text, such as physician notes and discharge summaries, is not only more readily available but also contains nuanced, context-rich information that structured data often lacks^19^.

Surprisingly, signal data, despite its clinical relevance and frequent use in practice (e.g., EKG in cardiology, EEG in neurology, Doppler in radiology), was underrepresented in the studies reviewed (n=16). This underrepresentation may stem from the challenges associated with digitizing, collecting, and processing signal data in healthcare settings - e.g., most of the time, EKG results are on paper. Similarly, omics data, which hold immense promise for personalized medicine, were rarely employed, likely due to their complexity, high dimensionality, and the cost associated with their acquisition and integration^20^. These gaps highlight opportunities for future research to explore the potential of these underutilized modalities.

Another notable finding pertains to the predominance of late fusion methods in multimodal AI development. In contrast, early fusion was rare, as it is unsuitable for many complex data modalities. While late fusion is often favored for its simplicity and modularity, it comes with the potential drawback of missing interactions between modalities that could yield valuable insights. Hybrid or joint fusion was also infrequently used, probably due to its complexity^21^. However, as frameworks supporting these joint fusion strategies continue to evolve, their adoption is expected to increase, offering a more nuanced approach to capturing cross-modal interactions.

While this systematic review highlights the superiority of multimodal approaches over unimodal algorithms in most studies, these findings should be interpreted with caution due to several critical limitations and uncertainties.

The evaluation of machine learning algorithms in healthcare is not standardized in the retrieved articles. Studies employed a wide range of metrics (e.g., AUC, C-index, confusion matrices). Establishing a standardized framework for evaluation metrics tailored to task-specific requirements is essential to advance the field^22^.

A notable issue in the reviewed studies is the limited use of proper validation datasets, which undermines the trustworthiness of the reported results. Studies that included a validation dataset often reported worse performance on the validation data compared to the test dataset, highlighting potential overfitting during model development. This emphasizes the need for robust validation protocols, including the use of external datasets^23^ and/or federated learning^24^, to ensure that results are generalizable and reliable in real-world clinical settings.

Publication bias may also influence the current understanding of multimodal machine learning. Studies reporting negative findings—such as no significant performance improvement with multimodality compared to unimodality—are probably less likely to be published, as seen in other areas of research^25^. Moreover, the prevalence of publication bias in this domain remains unclear. This bias skews the literature toward highlighting positive outcomes, potentially overstating the advantages of multimodal approaches.

The reasons behind the performance differences between multimodal and unimodal approaches remain unquantifiable. While clinical relevance of the added modality and the quality of the datasets (e.g., standardization and consistent ground truth labeling) likely play a role, the starting point might also play a significant role. For example, a unimodal algorithm with strong baseline performance (e.g., AUC > 0.95) may derive less benefit from multimodal integration than one with lower baseline performance. Interestingly, no correlation was found between sample size and added performance. The specific factors driving the gains of multimodality require further investigation.

Multimodal algorithms inherently complicate interpretability, as they often integrate multiple models, each processing a different data modality. While techniques such as SHAP can provide insights into the contribution of individual features within a single model, they do not extend easily across multiple interconnected models within a multimodal framework. As a result, understanding how different modalities interact and contribute to the final prediction becomes more challenging. Despite the importance of explainability in healthcare, few studies explicitly addressed this issue. Moreover, handling bias in multimodal data presents unique challenges^26^, as the integration of modalities with differing biases or noise levels can propagate or exacerbate existing biases, further complicating model validation and clinical adoption^27^.

At last, the gap between academic research and clinical practice remains significant. None of the studies included a medico-economic evaluation, and only one study referenced regulatory clearance. Additionally, only a small fraction of studies provided open-source algorithms, limiting reproducibility and broader accessibility. These gaps highlight the need for greater emphasis on translational research, ensuring that advances in multimodal machine learning can be effectively and ethically integrated into clinical workflows.

## Limits

This systematic review is not without limitations, many of which stem from the inherent complexity and heterogeneity of the subject matter. Multimodal machine learning in healthcare is a broad and rapidly evolving field, and the scope of the literature examined reflects this diversity. The included studies varied widely in terms of clinical decision-making tasks, data modalities, algorithmic approaches, and evaluation methods, making direct comparisons challenging.

Therefore, data extraction posed additional challenges due to the multifaceted nature of many articles. Several studies addressed multiple clinical decision-making tasks or employed a variety of algorithmic approaches and evaluation methods within the same publication. In such cases, we selected the approach most prominently highlighted in the article, often relying on the abstract as a guide. While this approach ensured consistency, it may have introduced bias, as certain methods or results emphasized in the article may not fully represent the breadth of experiments conducted. This selection process could have influenced our analysis and interpretation.

Another limitation relates to the potential biases inherent in the systematic review process itself. Despite efforts to conduct a comprehensive literature search, it is possible that relevant articles were missed due to the search strategy or database limitations. Additionally, while we employed the PROBAST framework to assess the risk of bias in the included studies, it is important to acknowledge that PROBAST was originally designed for traditional prognostic and diagnostic models and was not specifically developed for multimodal machine learning. As a result, certain methodological challenges unique to multimodal approaches—such as the handling of missing data across different modalities, the integration of heterogeneous sources, and the potential for compounding biases—may not have been fully captured by this assessment.

We acknowledge that some papers evaluating multimodal algorithms might be missed because they do not use the exact “multimodal” keyword. However, these papers do not compare the multimodal approach to unimodal approaches as it is not their aim.

Finally, this review underscores a significant challenge in the field of machine learning: the issues of reproducibility and replicability, exacerbated by the frequent lack of accessible data and code^28^. Reproducibility—the ability to achieve consistent results using the same data and methods—and replicability—the capacity to obtain similar outcomes with new data—are foundational to the modern scientific method. However, in machine learning research, these principles might sometimes be compromised. For instance, concerns have been raised that machine learning could be fueling a reproducibility crisis in science^29^. Similarly, the reproducibility issues in healthcare AI have been documented, emphasizing the need for accessible data and code to validate models effectively^30^. Addressing these challenges necessitates a cultural shift towards open science practices, where researchers routinely share their data and code, thereby enhancing the credibility and utility of machine learning applications in healthcare.

Only one previous pre-print review has examined the added performance of multimodal machine learning in healthcare^31^. Nevertheless, it differs fundamentally from ours in their scope and objectives. Unlike our study, which focuses exclusively on studies that explicitly compare unimodal approaches to multimodal approaches, these reviews included a much broader range of multimodal AI research without systematically evaluating the added performance of multimodality over unimodality. This explains why we did not include some studies that assess the relevance of multimodal LLM in healthcare: these papers do not compare multimodal LLM to unimodal ones. Furthermore, while their scope spanned various applications, we restricted our analysis solely to three very specific clinical decision-making tasks. This much narrower focus allows our review to directly address the key question of whether multimodality provides a meaningful performance advantage for clinical decision support, an issue that remains crucial for the translation of multimodal AI into real-world healthcare settings. Also, this paper did not assess the risk of bias of the studies included.

Despite these limitations, this systematic review provides valuable insights into the current state of multimodal machine learning in healthcare and identifies critical gaps and opportunities for future research. Addressing these limitations in subsequent studies and reviews will be essential for advancing the field.

## Conclusion

In the training of clinical decision-making algorithms, multimodal approaches demonstrated superior performance compared to unimodal data in 87% of included studies, with a potential benefit towards the combination of machine learning and deep learning approaches, compared to the deep learning only methodology. However, this finding should be interpreted with caution due to methodological heterogeneity and publication bias. Future research is needed to clarify specifically the relevance of evaluation metrics and hybridation model architecture depending on the clinical task.

## Methods

### Literature review

A systematic review of academic papers published from inception to January 2025 was performed.

This systematic review follows Preferred Reporting Items for Systematic Reviews and Meta-Analyses [PRISMA] 2020 recommendations^32^, which describes an evidence-based minimum set of items for systematic review reporting and diagnostic study meta-analyses, and is registered under the number CRD420250651820 (PROSPERO, https://www.crd.york.ac.uk/prospero/), ensuring that the review methods were established prior to the conduct of the review.

### Search Strategy

In January 2025, the PubMed (MEDLINE) database was queried to identify potentially relevant articles using the below search strategy and inclusion/exclusion criteria (**Table 4**).

**Table 4:** Inclusion and exclusion criteria.

| Inclusion criteria | Exclusion criteria |
| --- | --- |
| Peer-reviewed original research articles | Studies that does not fulfill all the inclusion criteria |
| Research conducted in consent humans above 18 years old | Study that are not original research (e.g. review, meta-analysis, editorial, et cætera.) |
| Compare the performance of multimodal AI to unimodal AI | Some parts of the article is inaccessible, e.g. supplementaries, even after contacting the authors |
| Explicitly involve the use of multimodal AI, integrating at least two different types of data (e.g., imaging and genetic information, clinical notes and laboratory results) between tabular data, free text, images, videos, signal and omics |  |
| Across any field of medicine |  |
| Across any clinical decision-making: diagnosis, prognosis, therapeutic decision, etc. |  |
| Algorithm relevance as a primary outcome, measured in terms of metrics such as sensitivity, specificity, positive predictive value, negative predictive value, precision, or area under the receiver operating characteristic curve (AUC) |  |
| Studies utilizing real patient data, rather than purely synthetic or simulated datasets, to ensure the clinical applicability of the findings |  |
| Include articles published in English, French, Spanish, Chinese or Arabic |  |
| Studies must have a clearly described methodology for AI model development, including data sources, model training, validation, and testing processes, to ensure methodological transparency and reproducibility |  |
| Studies should mention ethical approval or exemption where applicable, ensuring ethical standards in research involving human participants. |  |

To proceed with the screening, all searched articles were imported into the Rayyan tool^33^. Three reviewers (AB, SO and FL) independently screened each title and abstract. As the inclusion and exclusion criteria were quite specific, we did not perform a title-only screening - the relevant information being frequently on the abstract. The conflicts were resolved with the help of a fourth and a fifth author (PB and XT). The authors (SO, PB, FL and AB) then performed a full-text analysis for assessment. Conflicts were then resolved through consensus with one additional author (XT). The authors (SO, PB and AB) then extracted features of the selected articles.

### Search Query

The request was performed on 27th January 2025 (**Figure 5**). It is composed of three main parts: the first one aims to capture articles dealing with multimodal artificial intelligence (Title and Abstract), the second one aims to capture articles that evaluate algorithms through classic metrics (Full text) and the third one aims to limit the articles to the medical scope (Full text). For each part, we included as many synonyms as we identified. We did not include MeSH terms as we acknowledge that papers with an informatics background do not often include MeSH terms.

**Figure 5:**
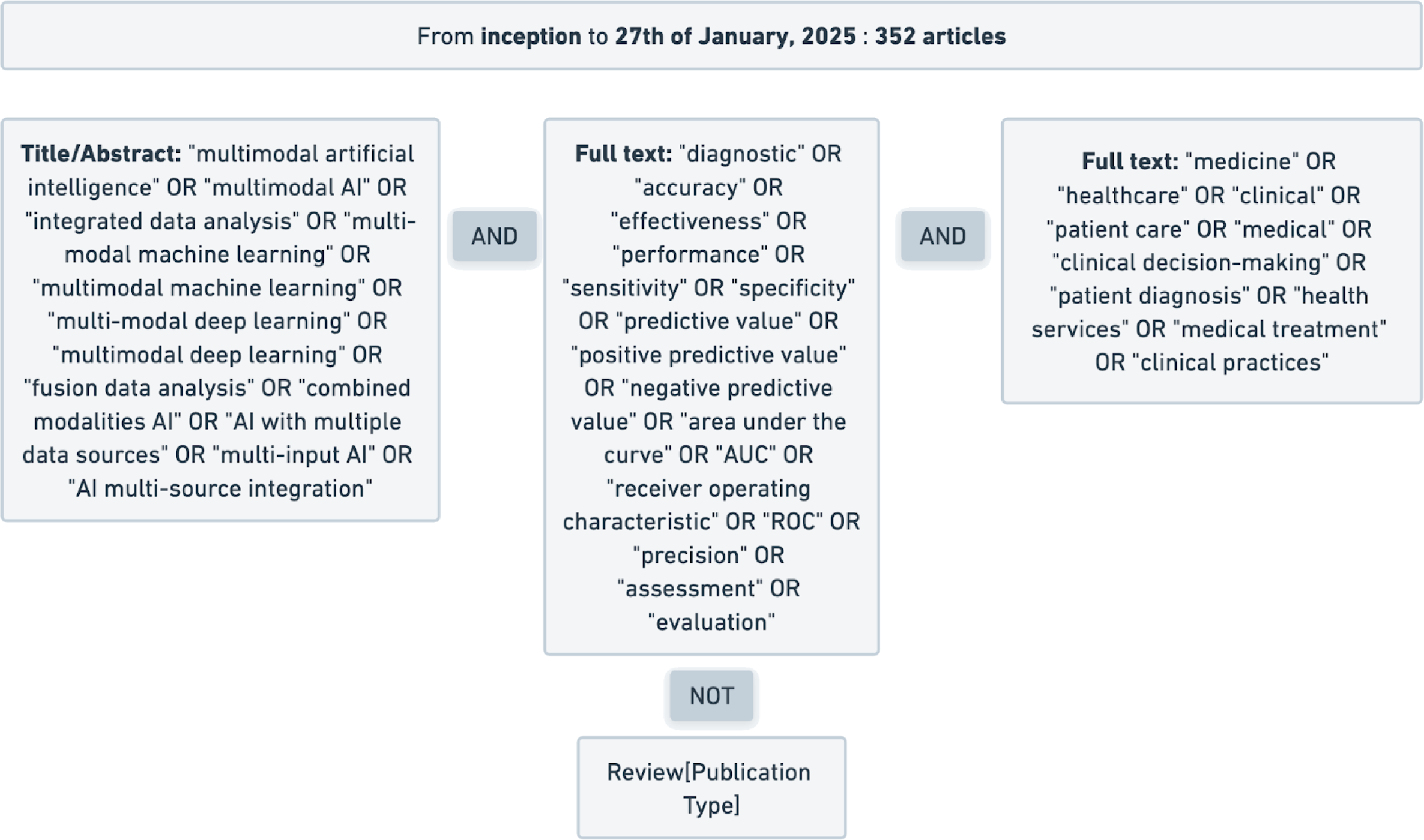
Search strategy. Search string visualisation. Visualisation of the keywords and logical connections that formed the search string. Each box can be translated to parentheses in the search string. Keywords inside each box are connected with each other by a logical OR. Unless indicated otherwise, all keywords are focused on Title and Abstract only.

We excluded review articles at the request phase. Despite that, some reviews were retrieved and were excluded in subsequent phases.

### Eligibility Criteria

We included peer-reviewed original research articles, of any design, across any field of medicine, that explicitly compared a multimodal algorithm to a unimodal algorithm for a clinical decision making task (diagnosis, prognosis or prescription). We did not include studies that compared multimodality to other set-ups than unimodal algorithms, like physician performances, as it is not in the scope of this systematic review. Studies must have a clearly described methodology for AI model development, including data sources, model training, fusion methodology, and model validation, to ensure methodological transparency and reproducibility. We only included studies which explicitly compared unimodal strategy to multimodal strategies. They must have compared the two strategies in a bias-controlled way as much as possible, ensuring that all aspects of the study remain equal beside the modalities. All duplicates were removed. A duplicate is defined as an article led by the same team, using exactly the same multimodal algorithm for the same task on the same databases.

### Data Extraction Strategy

A predesigned data collection form was prepared to extract the relevant information from the selected studies, including the title, date of publication, name of the journal, list of authors, keywords used, conditions tackled, outcome measured, modalities used, origin of the population, total available data, train sample size, test sample size, database centralization, evaluation methodology, algorithms used in the unimodality settings, algorithms used in the multimodality settings, fusion methodology, evaluation metric, unimodality best score, multimodality best score, license of the algorithms, MD regulation process status, medico-economic evaluation, and report of a competing interest.

The score extracted was the one most highlighted in the article. An evaluation metric was considered emphasized if it was highlighted in the article abstract and/or presented as the primary metric deemed most relevant and appropriate for the clinical use case under investigation. However, when the test set score was available, we extracted that one. When details were provided, we extracted the performance that seems to give the unimodal and multimodal approach the best chances. For the comparison to multiple unimodal methods, we extracted the modality with the best score.

To ensure comparison between the articles, we extracted as much as possible the AUC as the metric of reference, as it was the metric most frequently used in the articles included in this study.

Additionally, an analysis of the risk of bias was conducted following the PROBAST guidelines.

During the preparation of this work, we used ChatGPT (version 4o-2024-08-06 and GPT-4o1, OpenAI) to optimize the readability, feature extraction process, and wording of the manuscript. After using this tool, the authors reviewed, edited and completed the full content. The authors take full responsibility for the content of the publication.

## Author contribution

AB queried the database. AB, SO, PB and FL screened articles, extracted the data, performed statistical analyses and wrote the manuscript. EM, SB, XT supervised the methodological approaches and were major contributors in writing the manuscript. All authors read and approved the final manuscript.

## Data and code availability

All the data used in this paper is available <u>here</u> : https://bit.ly/tables_figures. The code used for the figures in this paper is available <u>here</u> : https://bit.ly/git link.

## Data Availability

All the data used in this paper is available here : https://bit.ly/tables_figures
The code used for the figures in this paper is available : https://bit.ly/git__link

https://bit.ly/git__link

## Acknowledgments

Authors thank Paul Roujansky, Nassim Hmidou, Damien Grosgeorge, Wilfried de Kerchove de Denterghem, Guillaume Goudot, Tristan Mirault, Lina Khider, and Alexandre Hollard for their technical and medical advices during this work.

## Conflict of Interest

AB, SO and PB are Zoī employees. All authors declare no conflict of interest regarding the present work.

## Abbreviations

AI: artificial intelligence
AUC: area under the curve
C-Index: concordance index
CT: computed tomography
EKG: electrocardiogram
GAN: generative-adversarial network
IF: impact factor
LLM: large language model
ML: machine learning
MRI: magnetic resonance imaging
NN: neural network
RF: random forest
SHAP: SHapley Additive exPlanations
SVM: support vector machine
US: ultrasound

## Funding

Some authors (AB, SO and PB) are employees of Zoī. Zoī had no implication in the methodology nor the writing of the paper.

## Notes

### Clinical Protocols

https://www.crd.york.ac.uk/PROSPERO/view/CRD420250651820

### Funding Statement

Some authors (AB, SO and PB) are employees of Zoi. Zoi had no implication in the methodology nor the writing of the paper.

