## Supplements for "Is Multimodal Better? A Systematic Review of Multimodal *versus* Unimodal Machine Learning in Clinical Decision-Making"

* Authors have the same involvement.

(1) Preventive Medicine, Data Science and AI Lab, Zoī, F-75010, Paris, France
(2) Limics INSERM U1142, Sorbonne Université, Université Sorbonne Paris-Nord, F-75006 Paris, France
(3) Département cardio-vasculaire, hôpital européen Georges-Pompidou, université Paris Cité, Inserm UMR 970 (équipe 2), 20, rue Leblanc, F-75015 Paris, France.
(4) Department of Women and Children's Health, St Thomas' Hospital, King's College London, London, UK
(5) Université de Paris Cité, AP-HP, Hôpital Universitaire Necker Enfants Malades, Service d'Imagerie Adulte, F-75015, Paris, France
(6) Sorbonne Université, CNRS UMR 7371, INSERM U 1146, Laboratoire d'Imagerie Biomédicale (LIB), F-75006, Paris, France
(7) Center for Transplantation Sciences, Massachusetts General Hospital, Harvard Medical School, Boston, Massachusetts, USA

**Abstract :**

Machine learning has demonstrated success in clinical decision-making, yet the added value of multimodal approaches over unimodal models remains unclear. This systematic review evaluates studies comparing multimodal and unimodal ML algorithms for diagnosis, prognosis, or prescription. A comprehensive search of MEDLINE up to January 2025 identified 97 studies across 12 medical specialties, with oncology being the most represented. The most common data fusion involved tabular data and images (67%). A risk of bias assessment using PROBAST revealed that 57% of studies had a low risk of bias, while 41% had a high risk. Multimodality outperformed unimodality in 91% cases. No correlation between dataset sample size and added performance has been observed. However, considerable methodological heterogeneity and potential publication bias warrant caution in interpretation. Further research is needed to refine evaluation metrics and hybrid model architectures based on specific clinical tasks.

Supplementary files

**Supplementary table 1 :  Articles screened during the full-text phase**

link

**Supplementary table 2 :  Centers characteristics of included studies**

link

| Type | Number | % |
|---|---|---|
| Multicentric | 51 | 51,52% |
| Monocentric | 39 | 39,39% |
| Bicentric | 9 | 9,09% |
| Total | 99 | 100,00% |

**Supplementary table 3 :  Distribution of year of publication**

link

| Year | Count | % |
|---|---|---|
| 2017 | 2 | 2,02% |
| 2019 | 3 | 3,03% |
| 2020 | 4 | 4,04% |
| 2021 | 10 | 10,10% |
| 2022 | 12 | 12,12% |
| 2023 | 22 | 22,22% |
| 2024 | 36 | 36,36% |
| 01/2025 | 10 | 10,10% |
| Total | 99 | 89,90% |

**Supplementary table 4 : Correlation between clinical significance of added modality and added performance**

link

| CONTINGENCY TABLE | delta above or equal (2) | delta below (1) | total |
|---|---|---|---|
| meaning high (3) | 22 | 21 | 43 |
| meaning moderate (2) | 12 | 9 | 21 |
| meaning low (1) | 0 | 2 | 2 |
| total | 34 | 32 | 66 |

**Supplementary figure 1 : Geographic distribution of datasets**

link

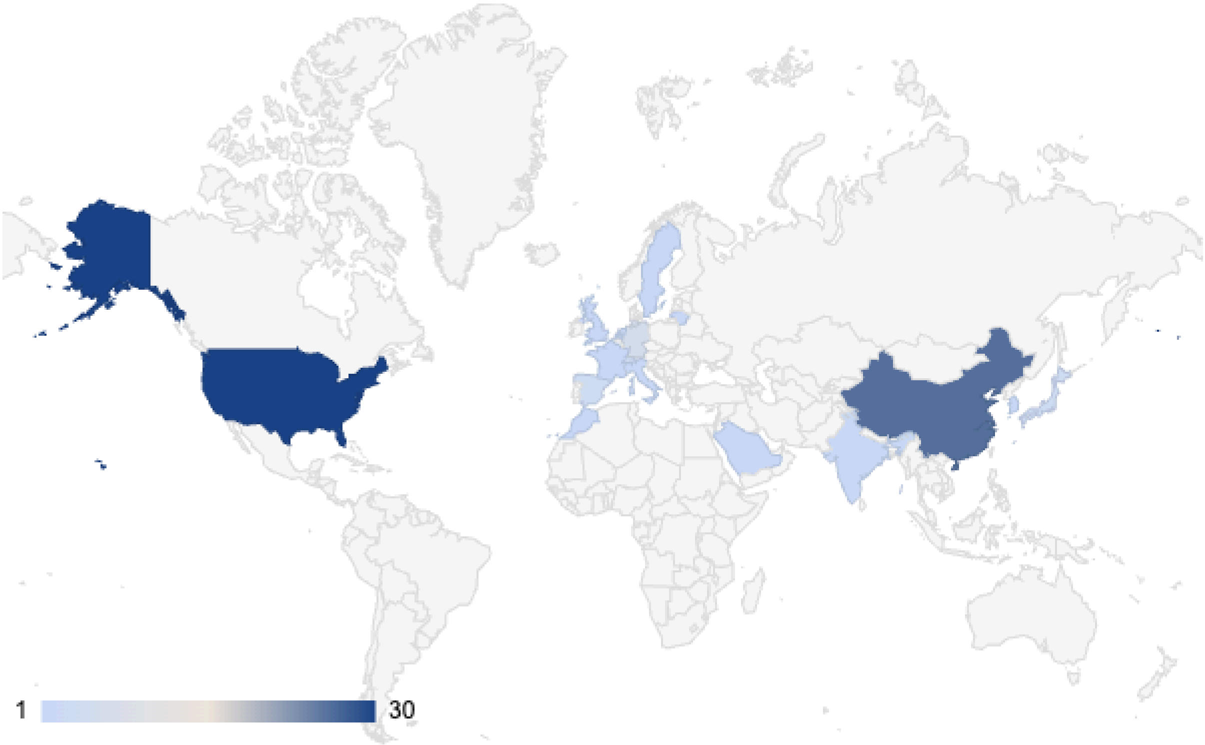

**Supplementary figure 2 : Distribution of medical specialities**

link

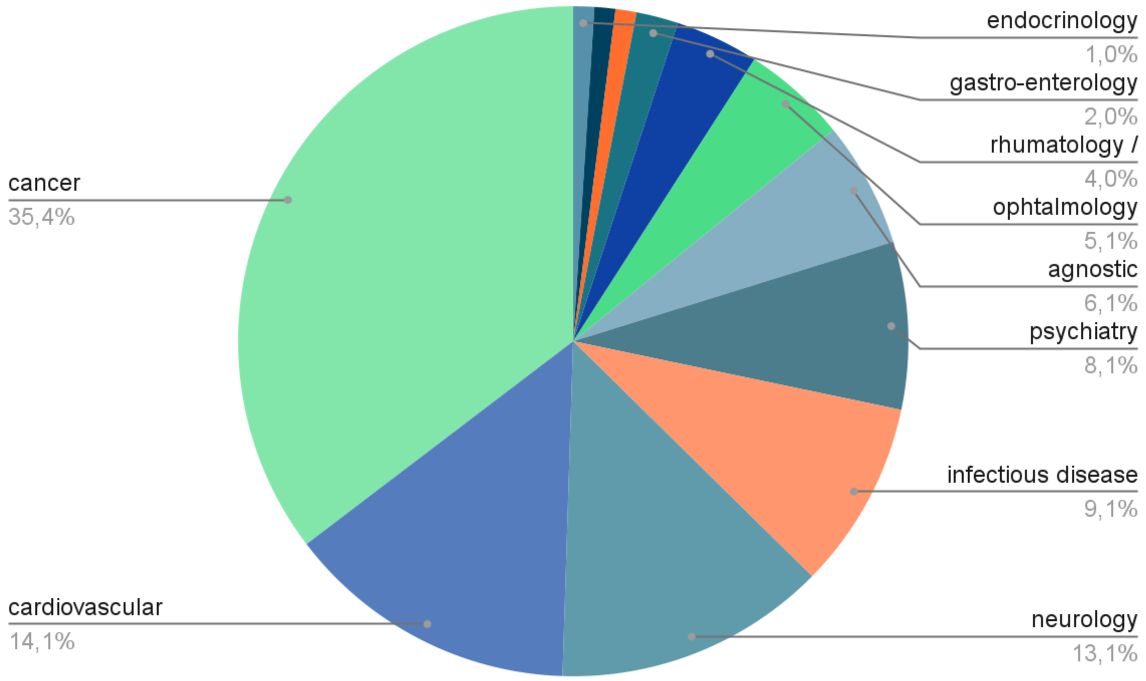

**Supplementary figure 3 : Distribution of algorithms tasks**

link

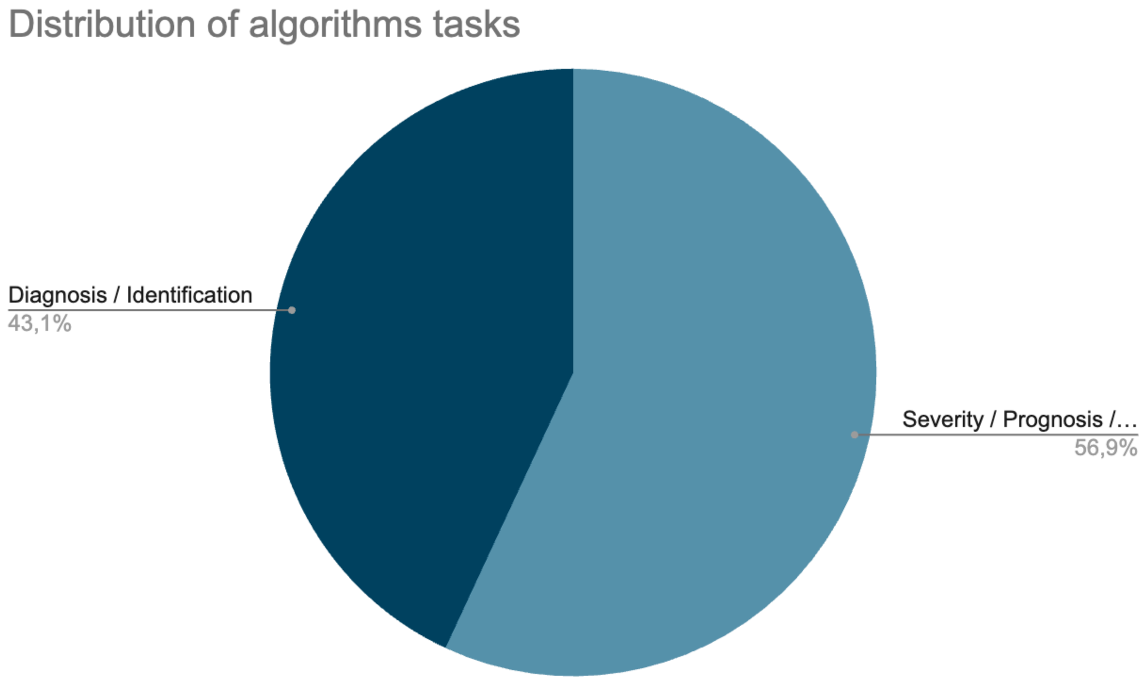

**Supplementary figure 4 : Distribution of evaluation metrics**

link

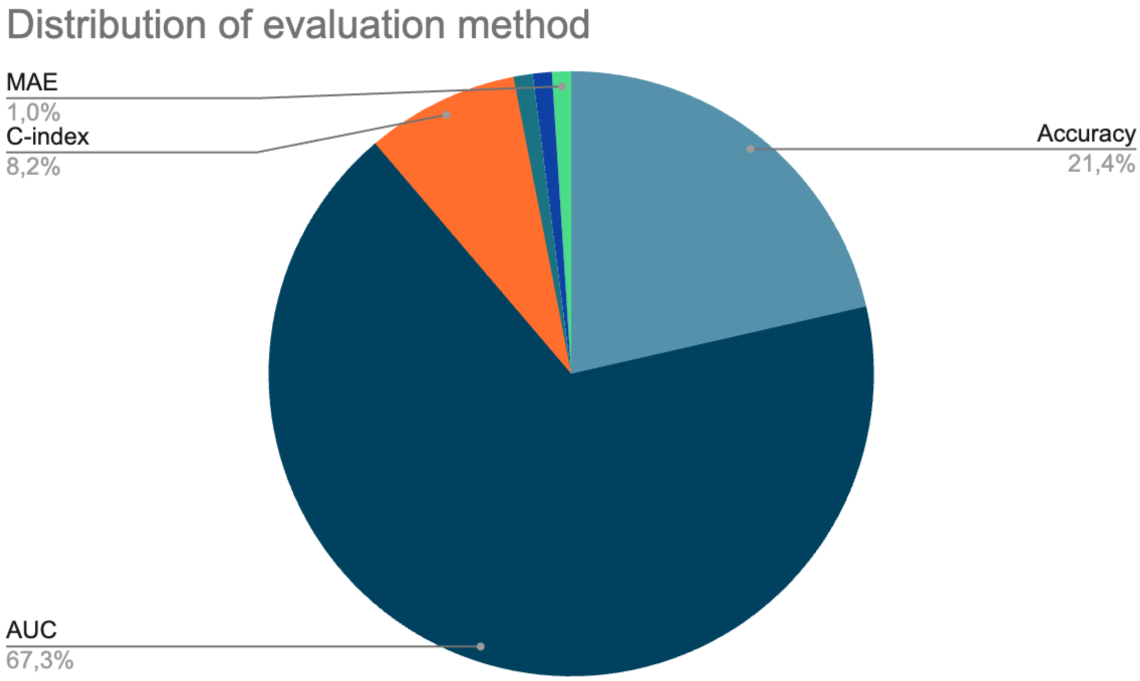
